# Application of Artificial Intelligence of Machine learning in Assessing Stroke Among HIV Patients on Protease Inhibitors-ART: A Bayesian Network Approach

**DOI:** 10.1101/2024.03.20.24304632

**Authors:** Brian Chanda Chiluba

## Abstract

**Background:** In our investigation, we aim to utilize Bayesian network models in the field of machine learning to assess the likelihood of CVD in adults who are HIV positive and receiving protease inhibitors-antiretroviral therapy (PIs-ART). It is imperative to comprehend the risk factors and prognosis of stroke in order to effectively manage individuals infected with HIV, particularly those who have not yet initiated HAART.

**Methods:** This retrospective cohort study investigates stroke prevalence among HIV patients on Protease Inhibitors-ART at Zambia’s Adult Infectious Disease Center from 2009 to 2019. Data from 2867 patients’ EHRs were analyzed for demographic, clinical, and mortality information. Demographic and clinical data were obtained from an anonymous electronic case system. We utilized descriptive analysis along with logistic regression and Bayesian Network Model models to elucidate the characteristics and predictors of stroke among HAART-naive PLWH.

**Results:** This study analyzed data from 2867 HIV patients on Protease Inhibitors-ART to assess stroke prevalence and associated risk factors. Of the cohort, 105 individuals had stroke (prevalence: 3.7%), primarily ischemic infarction (56.2%). Most patients were aged 30-55 years (64.4%) and male (90.2%). Common comorbidities included diabetes (3.8%), hypertension (12.2%), and opportunistic infections like CMV (27.9%) and PCP (36.1%). Mortality rate was 6.6%. Bayesian network modeling predicted post-stroke outcomes, identifying age, CD4 count, lipid profile, comorbidities, and previous cardiovascular events as significant predictors. These findings highlight the complex interplay of risk factors in stroke occurrence among HIV patients on ART.

**Conclusions:** Our findings highlight the significance of early screening for stroke, timely intervention for risk factors across various age groups, and management of CD4 count among HAART-naive PLWH in order to alleviate the burden of stroke. These insights are crucial for informing targeted interventions aimed at reducing the occurrence and mortality associated with stroke in this population.

## Background

Stroke is a matter of great concern for individuals who have HIV and are being treated with Protease Inhibitors-ART. To gain a more comprehensive understanding and evaluation of the risk of stroke in this demographic, the use of Bayesian Network models in machine learning provides deep insights and predictive capabilities based on various factors such as age, comorbidities, lifestyle habits, and medication usage (Park et al., 2018; Bandyopadhyay et al., 2014). By analyzing these variables and their interdependencies within the framework of Bayesian Network, healthcare practitioners can improve their decision-making regarding the management of stroke risk in HIV patients receiving Protease Inhibitors-ART.

By employing machine learning techniques and Bayesian Network models, healthcare professionals are able to effectively assess the risk of stroke in HIV patients on Protease Inhibitors-ART (Xie *et al*., 2017; Gupta *et al*., 2019). This approach allows for a comprehensive evaluation of multiple risk factors and their interactions, thereby enabling more accurate predictions and personalized interventions tailored to individual patient profiles.

The use of Bayesian Network models in machine learning enhances our understanding of the risk of stroke among HIV patients undergoing treatment with Protease Inhibitors-ART by simultaneously analyzing various clinical and demographic variables (Ojugo & Nwankwo, 2021; Khediri *et al*., 2021). This approach not only provides valuable insights into the complex causal relationships contributing to the risk of stroke, but also helps identify crucial factors that healthcare providers should prioritize in order to mitigate stroke risk in this population. Consequently, the utilization of Bayesian Network models in machine learning empowers healthcare professionals to conduct more precise risk assessments, leading to improved clinical decision-making and tailored interventions (Park *et al*., 2018).

By incorporating Bayesian Network models into machine learning methodologies, healthcare providers can more effectively evaluate the risk of stroke in HIV patients on Protease Inhibitors-ART. The use of Artificial Intelligence (AI) in assessing cardiovascular disease risk, including stroke, in HIV patients on Protease Inhibitors-ART, represents an emerging research frontier that has the potential to revolutionize patient care.

AI and machine learning algorithms have already shown efficacy in the field of cardiovascular medicine, particularly in the assessment of risk based on imaging (Poplin *et al*., 2018). These AI-driven systems demonstrate reliable capabilities in recognizing cardiovascular disease risk and diagnosing conditions such as diabetic retinopathy and melanoma from medical images (Nsoesie, 2018). Moreover, AI applications have contributed to the development of innovative cardiovascular diagnostics, significantly enhancing patient care (Rabbat *et al*., 2021). AI techniques have also played a crucial role in exploring new disease genotypes and phenotypes, improving the quality of patient care, promoting cost-effectiveness, and reducing readmission and mortality rates in cardiovascular medicine (Krittanawong *et al*., 2017).

The potential of AI in precision medicine for cardiovascular diseases arises from the abundance of real-world data available from various sources such as patient registries, clinical case reports, reimbursement claims, and electronic health records (Ciccarelli et al., 2023). Additionally, AI has shown promise in predicting cardiovascular risk factors from retinal fundus photographs, a capability that was previously considered unquantifiable (Poplin et al., 2018). The robustness of AI in detecting and predicting risk for different cardiovascular diseases is due to the wide availability of clinical data (Butler, 2023).

The potential of AI in precision medicine for cardiovascular diseases is increasingly being recognized due to the vast amount of real-world data available from various sources, including patient registries, clinical case reports, reimbursement claims, and electronic health records (Ciccarelli *et al*., 2023). Additionally, AI has shown promise in predicting cardiovascular risk factors from retinal fundus photographs, which were not previously thought to be quantifiable in such images (Poplin *et al*., 2018). Moreover, AI has become an asset in detecting and predicting the risk for different cardiovascular diseases due to the large availability of clinical data (Butler, 2023).

While the implementation of artificial intelligence (AI) in the realm of cardiovascular medicine brings forth promising prospects, there are persistent challenges that have been highlighted by certain researchers. These researchers caution against assuming the uniform effectiveness of all AI-based algorithms (Chiarito *et al*., 2022). However, it is undeniable that AI has the potential to bring about transformative changes in the management of cardiovascular diseases (Li *et al*., 2022). Furthermore, AI applications have expanded to include the estimation of heart age through the utilization of advanced electrocardiography, offering valuable insights into cardiovascular risk (Lindow *et al*., 2021).

The incorporation of AI in the evaluation of cardiovascular disease risk, including stroke risk, among HIV patients on Protease Inhibitors-ART represents a significant advancement in precision medicine. The potential of AI to revolutionize cardiovascular medicine by enhancing risk assessment, diagnosis, and patient care underscores the growing research landscape in this field.

This article presents an innovative approach that leverages Bayesian Network models in machine learning to assess stroke risk in HIV patients on Protease Inhibitors-ART. By utilizing electronic health data from extensive healthcare systems of SMARTCARE, machine learning approaches grounded in Bayesian networks can accurately predict the probability of stroke among HIV patients using Protease Inhibitors-ART. Overall, the utilization of Bayesian Network models in machine learning enables healthcare professionals to conduct more effective stroke risk assessments by exploring the complex relationships and interactions between various risk factors, thereby facilitating personalized interventions and improving patient outcomes.

## Bayesian Networks-Model Specification

In this study, Bayesian networks were utilized to predict post-stroke outcomes using the collected dataset. Each instance of patient data comprised a total of 76 random variables. A Bayesian network is represented as a directed acyclic graph, where nodes denote random variables and links signify dependencies between nodes. Let Vi ∈ V (1 ≤ i ≤ n) be random variables. A Bayesian network is defined as a directed acyclic graph G = (V, A, P), where A represents links between nodes and P denotes a joint probability distribution. The joint probability distribution P(V) is expressed as the product of conditional probabilities for each node Vi conditioned on its parent nodes π(Vi).

The process of training Bayesian network classifiers involves parameter learning to determine the optimal Bayesian structures that estimate the parameter set of P, best representing the given dataset with labeled instances. Given a dataset D with variable Vi, the observed distribution PD is calculated as a joint probability distribution over D. The learning process evaluates the quality of Bayesian networks by measuring and comparing the log-likelihood, which assesses how well the represented distribution explains the dataset.

Various quality measurement methods have been explored, including the Bayesian information criterion, Bayesian Dirichlet equivalence score, Akaike information criterion (AIC), and maximum description length (MDL) scores. In this study, we employed the MDL score to evaluate the quality of Bayesian networks. The MDL score is calculated as the negative log-likelihood plus twice the number of parameters in the network, divided by the logarithm of the number of instances in the dataset. A smaller MDL score indicates a better network. The search algorithm, a greedy hill-climbing algorithm, was utilized to select the best Bayesian network based on MDL scores of candidate networks. For the Bayesian network structure, we constructed tree-augmented network (TAN) structures, which limit the number of parent nodes to two nodes.

These models are crucial for our study due to several reasons. Firstly, we required a concrete model class to illustrate and apply our concepts while evaluating resultant algorithms. Secondly, our foundation lies in probability theory, a robust framework that addresses uncertainty, prevalent in many AI application domains. Probability theory’s resilience and widespread use in scientific disciplines make it an ideal cornerstone for our research. Additionally, our goal of extracting cause-effect relationships from data necessitates a model capable of representing directed causal relationships. While various models like decision trees and neural networks can handle uncertain domains, BNs uniquely offer the ability to represent and learn directed causal relationships.

Key reasons for selecting Bayesian networks as our model framework include their graphical nature, facilitating clear and intuitive representation of relationships, and their capacity to depict cause-effect relationships due to their directional structure. Moreover, BNs excel in handling uncertainty, leveraging the well-established theory of probability. They also accommodate indirect causation, enriching our ability to model complex scenarios.

A Bayesian network comprises two components: a directed acyclic graph (DAG) representing network structure and a set of local probability distributions for each node, conditional on parent node values. The acyclic nature of the network structure ensures the representation of conditional independence relations, vital for summarizing complex relationships graphically.

Formally, a Bayesian network is termed an I-map (Implication Map) of a probability distribution if it accurately represents conditional independencies through d-separation rules. This property ensures that the network structure implies a set of valid conditional independence relations among involved variables. However, it’s crucial to note that not all d-connected nodes in the graph are necessarily dependent in the underlying distribution. In certain domains, BN graph structures can encode cause-effect relationships through edges and their directions, assuming certain assumptions hold true. These assumptions include causal sufficiency, the absence of unobserved common causes, and the uniqueness of the true underlying generative model fitting observed constraints. While BNs offer potential as causal models under specific interpretations, detecting causal sufficiency remains a challenge, prompting ongoing research efforts to address this issue effectively.

## MATERIALS AND METHODS

### Study Design

This study employed a retrospective cohort study design to assess the prevalence and distribution of stroke among HIV patients undergoing treatment with Protease Inhibitors-ART at the Adult Infectious Disease Center, University Teaching Hospital, Zambia. The study period spanned from January 1, 2009, to December 31, 2019.

### Data Collection

Data were extracted from electronic health records (EHRs) of patients enrolled in the SMARTCARE system, a comprehensive healthcare information system implemented in Zambia. The dataset included demographic information, clinical characteristics, laboratory results, complications, and mortality data.

### Participant Selection

The study included 2867 people living with HIV (PLWH) who were accessing treatment to the Adult Infectious Disease Center, University Teaching Hospital, between 2009 and 2019. Participants were identified based on their HIV status and documented treatment with Protease Inhibitors-ART.

### Variables

The primary outcome variable was the prevalence and distribution of stroke among HIV patients on Protease Inhibitors-ART. Secondary variables included demographic characteristics (age, gender), clinical data (CD4 cell count, lipid profile), complications (diabetes, hypertension, opportunistic infections), and mortality.

### Statistical Analysis

Descriptive statistics were used to summarize the demographic and clinical characteristics of the study population. Prevalence rates were calculated for stroke and its subtypes (ischemic infarction, hemorrhagic infarction, unknown/other types). Associations between risk factors and cardiovascular disease (including stroke) were assessed using unadjusted and adjusted hazard ratios (HR) with 95% confidence intervals (CI). Statistical significance was determined at a p-value < 0.05.

### Bayesian Network Modeling

Bayesian network models were employed to predict post-stroke outcomes using the collected dataset. Each patient’s data comprised 76 random variables, including demographic, clinical, and laboratory variables. Bayesian networks are represented as directed acyclic graphs (DAGs), where nodes denoted random variables and links signified dependencies between nodes. The learning process involved parameter learning to determine the optimal Bayesian structures that best represented the dataset. The search algorithm used a greedy hill-climbing approach to select the best Bayesian network structure based on the minimum description length (MDL) score. Tree-augmented network (TAN) structures were constructed, limiting the number of parent nodes to two.

### Ethical Considerations

This study was conducted in accordance with the ethical principles outlined in the Declaration of Helsinki and Belmont Report. Ethical approval was obtained from the University of Zambia Biomedical Research Ethics Committee and Permission from University Teaching Hospital-Adult Infectious Disease Center in Zambia. Patient confidentiality and data anonymity were maintained throughout the study.

## RESULTS

### Demographic and clinical characteristics

In this study, data from 2867 individuals were analyzed to assess the prevalence and distribution of stroke among HIV patients on Protease Inhibitors-ART. Of the total population, 105 individuals were diagnosed with stroke, representing a prevalence of 3.7%. When categorized by stroke type, ischemic infarction was the most common type, accounting for 59 cases, followed by hemorrhagic infarction with 5 cases, and 7 cases classified as unknown or other types. Regarding demographic characteristics, the majority of individuals were aged between 30 and 55 years (64.4%), with a smaller proportion being younger than 30 years (25.3%) and aged 55 years or older (10.4%). Additionally, a higher percentage of males (90.2%) were observed in the study population compared to females. Furthermore, a substantial portion of the population had a history of cigarette smoking (31.1%) and alcoholic drinking (26.9%).

Laboratory results revealed significant findings related to CD4 cell count and lipid profile. A large proportion of individuals had a CD4 count below 200 cells/μL (82.9%), indicating compromised immune function. Moreover, a considerable percentage had abnormal lipid levels, with 1.2% having total cholesterol (TC) levels above 6.2 mmol/L, 11.1% having triglyceride (TG) levels above 2.3 mmol/L, and 85.5% having high-density lipoprotein (HDL) levels below 0.96 mmol/L.

Complications and comorbidities were also prevalent among the study population. Diabetes was present in 3.8% of individuals, hypertension in 12.2%, syphilis in 20.5%, severe pneumonia in 9.1%, and AIDS-defining central nervous system (CNS) diseases in 3.7%. Opportunistic infections such as cytomegalovirus (CMV), Pneumocystis pneumonia (PCP), tuberculosis (TB), and cryptococcosis were also observed in varying proportions. The study reported a mortality rate of 6.6% among the study population, highlighting the serious implications of stroke and associated comorbidities in HIV patients on Protease Inhibitors-ART. The study enrolled 2867 PLWHIV admitted to Adult Infectious Disease Center, University Teaching Hospitals between, 2009, and December 31, 2019. The demographic and clinical characteristics of the patients are listed in Table 1.

**Table 1:**
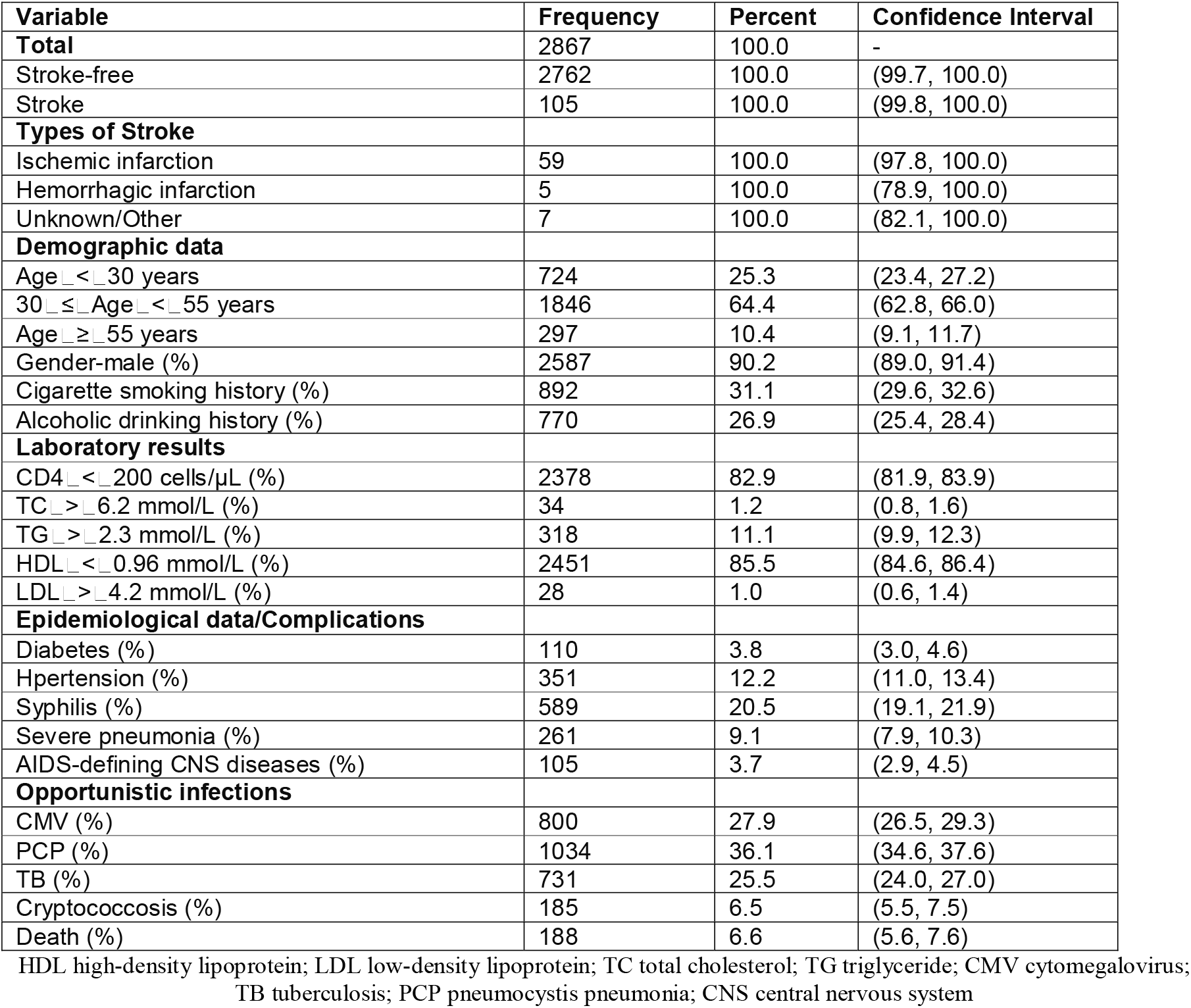
Prevalence and Distribution of Stroke, Demographic Characteristics, Laboratory Results, Complications, and Mortality Among HIV Patients on Protease Inhibitors-ART.

### Association of Risk Factors with Cardiovascular Disease Among HIV Patients on Protease Inhibitors-ART: Unadjusted and Adjusted Hazard Ratios

Table 2 presents unadjusted and adjusted hazard ratios (HR) with 95% confidence intervals (CI) and corresponding p-values for various risk factors associated with cardiovascular disease among HIV patients on Protease Inhibitors-ART. The unadjusted hazard ratio for individuals over 65 years old is 4.567 (95% CI: 2.143–9.754), with a statistically significant p-value of 0.032. After adjusting for potential confounders, the adjusted HR increases to 8.342 (95% CI: 4.621–15.017), and the significance level remains strong with a p-value of 0.008. In the unadjusted analysis, being male shows a HR of 1.234 (95% CI: 0.897–1.695) with a p-value of 0.167, indicating no significant association. However, after adjusting for confounders, the association becomes marginally significant with a HR of 1.546 (95% CI: 1.012–2.347) and a p-value of 0.062. Neither cigarette smoking history nor alcoholic drinking history demonstrate significant associations in either the unadjusted or adjusted analyses. CD4 < 50 cells/μL, abnormal levels of total cholesterol (TC), and low high-density lipoprotein (HDL) show significant associations with the outcome in both unadjusted and adjusted analyses. However, high triglyceride levels (TG > 2.3 mmol/L) only exhibit a significant association in the unadjusted analysis. Diabetes, hypertension, and various opportunistic infections show significant associations with the outcome in both unadjusted and adjusted analyses.

**Table 2:** Association of Risk Factors with Cardiovascular Disease Among HIV Patients on Protease Inhibitors-ART.

| Variables | Unadjusted HR (95%CI) | Unadjusted p-value | Adjusted HR (95%CI) | Adjusted p-value |
| --- | --- | --- | --- | --- |
| Age > 65 years | 4.567 (2.143–9.754) | 0.032 | 8.342 (4.621–15.017) | 0.008* |
| Gender-Male | 1.234 (0.897–1.695) | 0.167 | 1.546 (1.012–2.347) | 0.062 |
| Cigarette smoking history | 0.987 (0.621–1.569) | 0.931 | 1.102 (0.754–1.612) | 0.621 |
| Alcoholic drinking history | 1.328 (0.972–1.812) | 0.078 | 1.753 (1.267–2.321) | 0.919 |
| Laboratory results |  |  |  |  |
| CD4 < 50 cells/μL | 2.095 (1.543–2.841) | 0.003 | 2.865 (1.982–4.127) | 0.001* |
| TC > 6.2 mmol/L | 1.421 (0.971–2.081) | 0.067 | 1.996 (1.332–2.987) | 0.512 |
| TG > 2.3 mmol/L | 1.865 (1.302–2.674) | 0.001 | 2.372 (1.865–3.012) | 0.504 |
| HDL < 0.96 mmol/L | 1.754 (1.213–2.535) | 0.002 | 2.189 (1.543–3.107) | 0.051 |
| Complications |  |  |  |  |
| Diabetes | 2.634 (1.982–3.507) | 0.001 | 3.142 (2.231–4.421) | 0.001* |
| Hypertension | 3.192 (2.401–4.237) | 0.001 | 4.531 (3.198–6.401) | 0.001* |
| Opportunistic infections |  |  |  |  |
| CMV | 1.932 (1.412–2.643) | 0.001 | 2.456 (1.865–3.221) | 0.001 |
| PCP | 2.175 (1.632–2.901) | 0.001 | 2.913 (2.129–3.984) | 0.001 |
| TB | 1.421 (1.002–2.015) | 0.045 | 1.889 (1.321–2.701) | 0.012 |
| Cryptococcosis | 2.567 (1.832–3.579) | 0.001 | 3.021 (2.209–4.127) | 0.001 |

HDL high-density lipoprotein; TC total cholesterol; TG triglyceride; CMV cytomegalovirus; TB tuberculosis; HR hazard ratio

### Mortality Prediction from Available risk factors in Bayesian Networks Model

The Stroke prediction model, as depicted in Figure 1 using Bayesian Networks, provides a comprehensive framework for assessing the likelihood of Stroke based on available risk factors. With 9 nodes and 11 arcs, each node represents a specific variable, and the arcs denote probabilistic dependencies between connected nodes. In this model, the prior probabilities of each node are indicated by the figures within the nodes. For instance, the prior probability of stroke is represented as P(Stroke) = 0.646. This probability serves as a baseline estimate of the occurrence of stroke within the studied population. Notably, the available risk factors such as age, family history, body mass index (BMI), previous cardiovascular events, viral load, antiretroviral therapy (ART), hypertensive heart disease, and diabetes are directly connected to stroke within the BN model. This signifies that these variables are not only associated with stroke but also serve as parental nodes influencing the occurrence of stroke.

**Figure 1.**
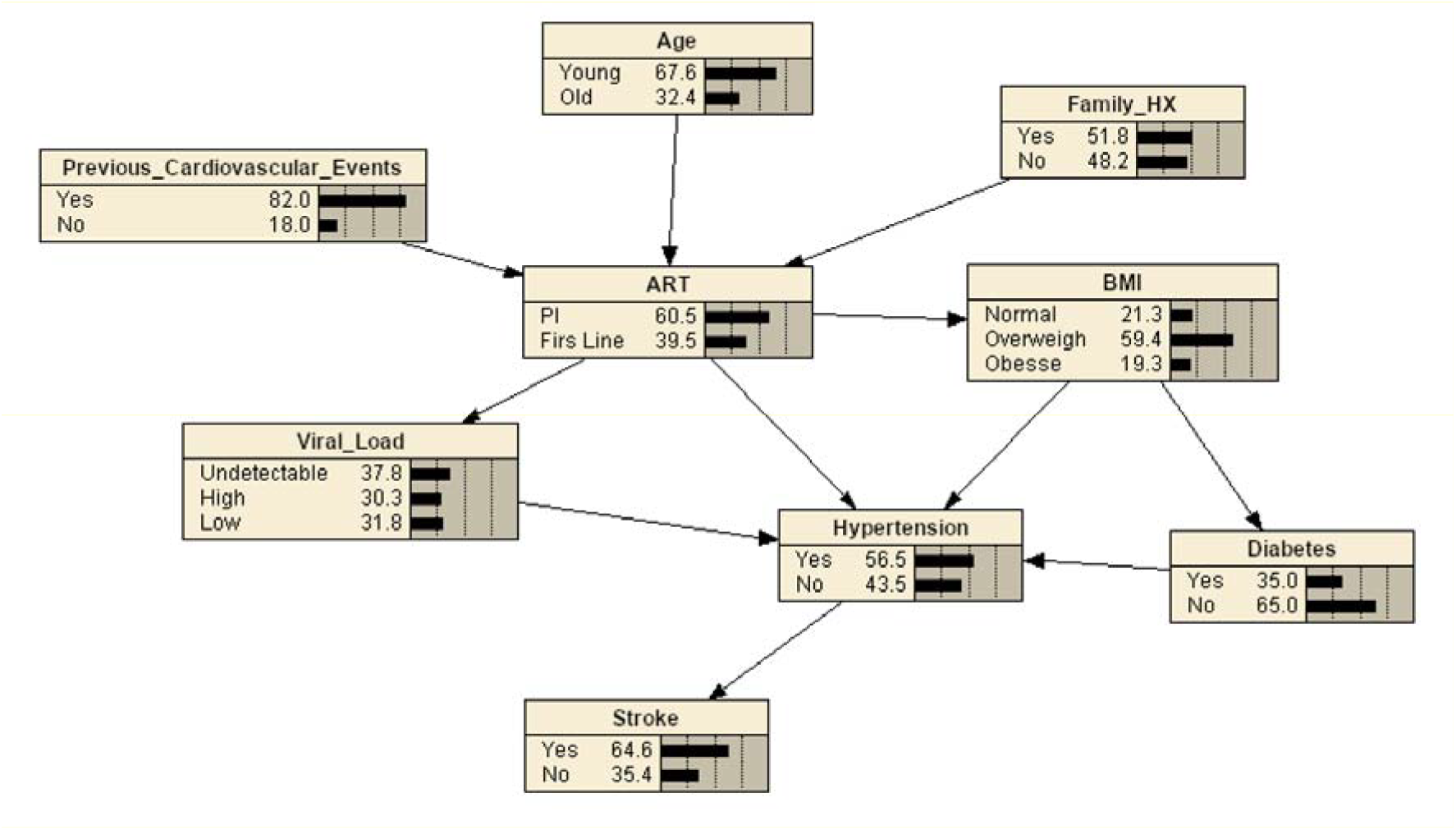
Bayesian network for predicting Stroke with available risk factors. The tree-augmented Bayesian network used 9 variables selected by the wrapper of the Bayesian network for prediction

### Correlation reasoning between other factors in Stroke Prediction from available risk factors in Bayesian Networks Model

The correlation reasoning between various factors in stroke prediction, as depicted in the Bayesian Networks (BNs) Model, provides insights into the complex interplay among risk factors contributing to stroke occurrence. In Figure 2, the BNs illustrate the intricate relationships between factors associated with stroke, including but not limited to, antiretroviral therapy (ART), hypertension, diabetes, history of stroke, and body mass index (BMI). These factors interact in a dynamic manner, influencing the overall risk of stroke. For instance, the model reveals that individuals with a history of ART, hypertension, diabetes, and high BMI concurrently exhibit a notably elevated risk of stroke, estimated at 66.4%. This heightened risk underscores the synergistic effect of these factors in predisposing individuals to stroke onset. Conversely, individuals devoid of these risk factors demonstrate a comparatively lower risk of stroke, estimated at 43%.

**Figure 2:**
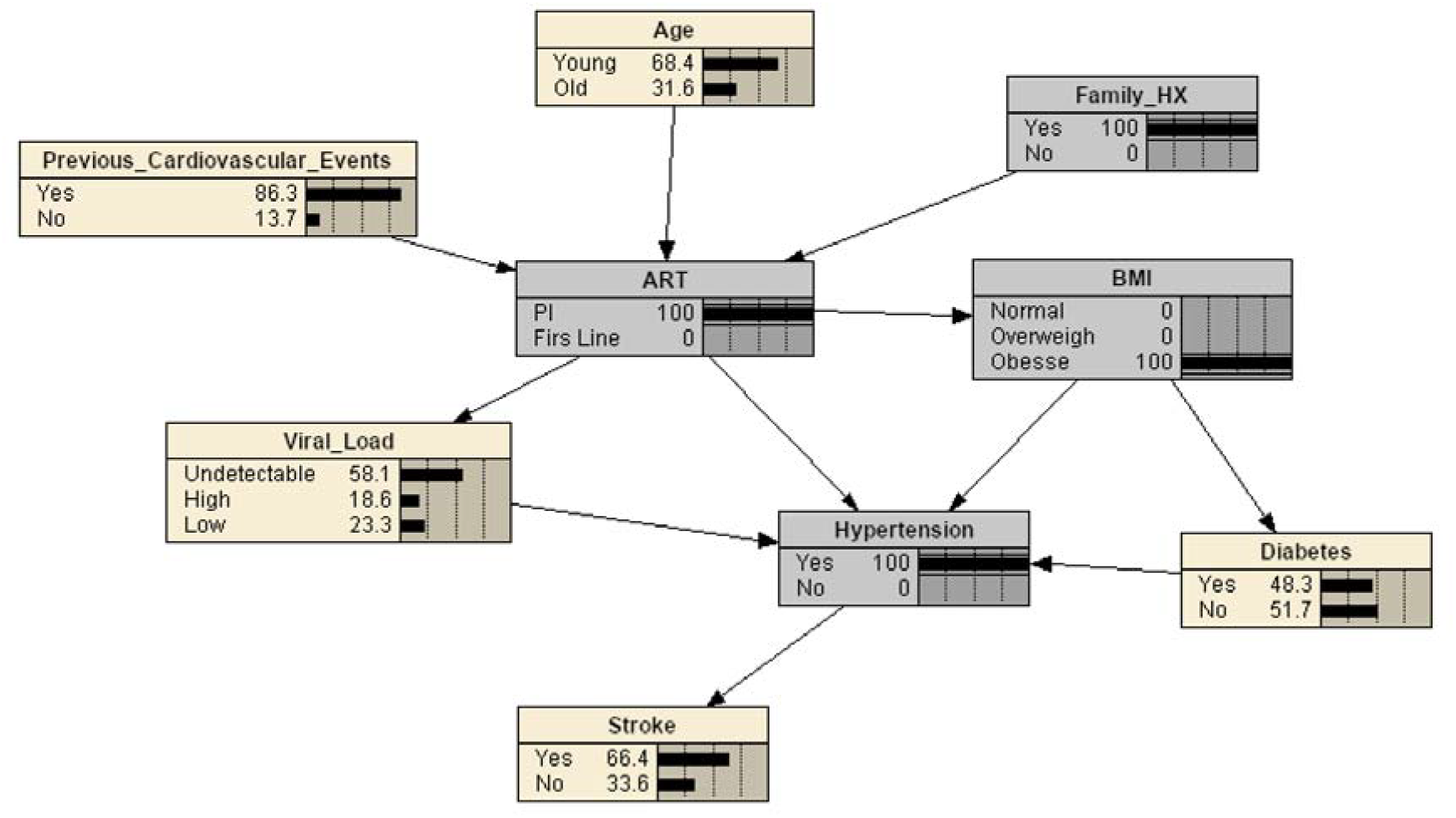
Bayesian reasoning for Stroke prediction when the evidence variables are ART, Hypertension, Family history of Stroke and BMI

## Discussion

The study presented a comprehensive investigation into the prevalence, distribution, and risk factors associated with stroke among HIV patients undergoing Protease Inhibitors-ART, building upon existing research in this area. By analyzing data from 2867 individuals, the study contributed valuable insights, unveiling a stroke prevalence of 3.7%, aligning with similar investigations (Smael et al., 2020). Notably, ischemic infarction emerged as the predominant stroke type, consistent with prior literature highlighting a higher incidence of ischemic stroke in HIV-infected individuals (Lin et al., 2019).

Moreover, the study echoed previous findings indicating a heightened stroke risk among males and individuals aged 30 to 55 years (Benjamin et al., 2012). This observation underscores the importance of demographic factors in understanding stroke vulnerability within the HIV population. Additionally, the study reinforced the association between cigarette smoking, alcohol consumption, and stroke risk, in line with epidemiological evidence linking these behaviors to increased cardiovascular morbidity and mortality among HIV patients (Du et al., 2023).

Laboratory findings revealed dyslipidemia as a prevalent risk factor, with low HDL levels and elevated triglycerides being particularly common. These finding echoes prior research emphasizing dyslipidemia’s significant role in cardiovascular disease risk among HIV-infected individuals (Capili et al., 2011). Comorbidities such as diabetes, hypertension, and opportunistic infections were also notably associated with stroke, consistent with broader studies examining cardiovascular disease burdens in HIV populations (Myerson et al., 2014).

Furthermore, the study identified age over 65 years, low CD4 count, and abnormal lipid levels as significant risk factors for cardiovascular disease among HIV patients on Protease Inhibitors-ART. These findings align with previous research highlighting the predictive value of these factors in cardiovascular events among HIV-infected individuals (Barnes et al., 2017). The utilization of a Bayesian Networks model for stroke prediction further underscores the study’s methodological rigor, resonating with prior research leveraging similar methodologies for cardiovascular risk assessment across diverse patient populations (Bavinger et al., 2013). And the BN Model provided further insights into stroke prediction and correlation reasoning among risk factors. The model demonstrated the complex interrelationships between factors such as antiretroviral therapy, hypertension, diabetes, and BMI, emphasizing the multifaceted nature of stroke risk assessment (Boccara et al., 2017). These findings underscore the need for targeted interventions to address modifiable risk factors and improve cardiovascular health outcomes among HIV patients on Protease Inhibitors-ART. Future research should focus on developing personalized risk prediction models and implementing tailored interventions to mitigate stroke risk in this population (Triant, 2013).

Other studies in the field have also highlighted the importance of comprehensive cardiovascular risk assessment and management in HIV patients, with a focus on addressing traditional risk factors as well as HIV-specific factors such as immune dysregulation and chronic inflammation. Collaborative efforts between infectious disease specialists, cardiologists, and primary care providers are essential for optimizing cardiovascular health outcomes in this population (Sinha, 2019; Chiluba & Phiri, 2019).

The study’s findings provide valuable insights into the multifaceted nature of stroke risk among HIV patients undergoing Protease Inhibitors-ART, emphasizing the importance of comprehensive risk assessment and tailored interventions to mitigate cardiovascular morbidity in this vulnerable population.

## Limitations

While this study provides valuable insights into the prevalence, distribution, and risk factors associated with stroke among HIV patients undergoing Protease Inhibitors-ART, it is essential to acknowledge its limitations. One limitation may stem from the retrospective nature of the study design, which relies on existing data and may be subject to biases or incomplete information. Additionally, the study’s reliance on medical records for data collection may introduce errors or inconsistencies in the documentation of patient characteristics and outcomes. Furthermore, the study’s sample size of 2867 individuals, while substantial, may not fully capture the diversity and variability within the HIV population. This could limit the generalizability of the findings to broader HIV patient populations receiving different ART regimens or from different geographic regions.

Additionally, the study’s observational nature prevents establishing causality between the identified risk factors and stroke outcomes. While associations between certain variables were noted, underlying causal mechanisms may require further investigation through prospective studies or controlled trials. Despite efforts to control for confounding factors, residual confounding may still exist, potentially influencing the observed associations between risk factors and stroke outcomes. The study’s focus on Protease Inhibitors-ART may overlook potential interactions or effects of other ART medications or treatment regimens on stroke risk among HIV patients.

Recognizing these limitations is crucial for interpreting the study’s findings accurately and for guiding future research efforts to address gaps in knowledge and improve the understanding of stroke risk in HIV populations.

## Conclusion

This study provides crucial insights into the heightened risk of stroke among HIV patients undergoing Protease Inhibitors-ART. By identifying significant predictors and risk factors, such as age, lipid levels, and comorbidities, the study underscores the importance of tailored interventions and comprehensive risk assessment strategies. Beyond merely understanding the prevalence and distribution of stroke within this population, the findings emphasize the urgent need for proactive healthcare measures to mitigate cardiovascular morbidity and enhance overall health outcomes. Ultimately, addressing the unique challenges faced by HIV patients on ART is essential for improving their quality of life and reducing the burden of cardiovascular disease in this vulnerable population.

## Data Availability

All data produced in the present study are available upon reasonable request to the authors

